# School-based social prescribing to address youth loneliness: a pilot randomised controlled trial

**DOI:** 10.64898/2026.04.27.26351665

**Authors:** D Hayes, J. Wright, F. Bu, N. Humphrey, P. Qualter, E. Han, L. Sticpewich, S. Maquire, L.C.G. Umpierrez, A. Burton, J. K. Bone, E. Stapley, M.S. Tibber, R. J. Booth, D. Fancourt

## Abstract

**Aims:** Youth loneliness is associated with poorer mental health, educational disengagement, and adverse long-term health outcomes. While social prescribing shows promise in reducing loneliness in adults, access for young people remains limited as most pathways sit within primary care. Schools offer a near-universal point of contact and may provide a more accessible referral route. The INACT study aimed to pilot a school-based social prescribing pathway and assess its feasibility, acceptability, and preliminary signals of benefit for loneliness and related outcomes.

**Methods:** Pupils in Years 4, 5, 7 and 8 (ages 9 to 13; n=672) across 11 schools in London, Manchester, and Leeds were screened for loneliness. Those reporting elevated loneliness (n=140, 20.8%) were randomised to social prescribing or signposting. For follow-up, the social prescribing arm was capped at 41 pupils, and a matched subset of the signposting group (n=45) was followed longitudinally. Outcomes were collected at baseline, three-, and six-months. Feasibility indicators included recruitment, uptake, engagement, and retention. Acceptability and appropriateness were assessed using validated implementation measures and qualitative interviews with pupils and link workers.

**Results:** One in five pupils reported elevated loneliness. In the social prescribing group, 83% initiated the pathway, attending a mean of 5.09 sessions, with most delivered in person. Implementation measures indicated high feasibility, acceptability, and appropriateness. Interviews highlighted the importance of relational support and personalised activity matching, although some younger pupils needed support completing questionnaires. Preliminary analyses showed reductions in loneliness at three-months in both groups, with somewhat greater reductions in the social prescribing arm.

**Conclusion:** School-based social prescribing appears feasible and acceptable, with early indications of benefit. Schools may offer scalable infrastructure for addressing youth loneliness beyond healthcare pathways. A fully powered multi-site trial with economic evaluation is now warranted to determine effectiveness and long-term value.

**Trial registration:** ClinicalTrials.gov: NCT06656663

## Introduction

Loneliness is increasingly recognised as a global public health concern associated with adverse mental and physical health outcomes across the life course^1^. Although loneliness has historically been framed as an issue primarily affecting older adults^2^, there is growing evidence that young people (YP) report some of the highest rates of loneliness^3^. In the UK, 11.3% of those aged 10–15 years report feeling lonely “often” or “always”^4^. Loneliness is socially patterned, with higher prevalence among younger individuals, those living in urban areas, and those from lower socioeconomic backgrounds^4^. If unaddressed, loneliness in childhood and adolescence is associated longitudinally with more depressive symptoms^5^, and has been linked to increased risk of cardiovascular disease and psychiatric disorders in adult-hood^6,7^. Proposed mechanisms include dysregulation of physiological systems involved in social connection and stress response^8^.

Recent meta-analyses reported a moderate overall effect of interventions on loneli-ness^9,10^, although interventions specifically targeting youth loneliness showed only small ef-fects^10^. Both meta-analyses also highlighted important evidence gaps, including limited targeting of YP experiencing loneliness and a need for interventions responsive to individual needs and contexts^9,10^. Research gaps were also identified, including (i) that interventions tended to target YP at risk of loneliness (e.g., due to health concerns), rather than those who reported being lonely, (ii) a lack of longer-term follow up, and (iii) a need to consider socio-economic and other risk factors in targeted interventions ^9,10^.

Social prescribing (SP) is a non-clinical pathway that links individuals to community-based support tailored to their interests, needs, and preferences^11^. It usually involves a health or allied health professional referring a patient to a Link Worker (LW), who co-develops a non-clinical plan based on what is important to the individual. Community support can vary, but could include art classes, sports, nature groups, volunteering opportunities, cultural activities, and broader advice and support services (e.g., debt or housing)^11^. SP aligns with calls for connector services that can increase social connections and improve relationships among people experiencing loneliness^12^.

Previous reviews suggest that SP may be an effective approach for tackling loneliness both in adults and YP^13,14^. However, methodological limitations regarding sample sizes and a lack of control groups mean caution is needed when interpreting study findings. More recently, research using robust designs with control groups has confirmed that SP can be beneficial in tackling loneliness^15,16^. In one UK matched comparator study (matched analyses n=613), 37% of adults receiving SP moved from being classified as lonely to not lonely at follow-up, compared with 20% of matched control^16^. There was also evidence that younger age groups benefitted more^16^. Similarly, a controlled trial in Australia exploring the impact of SP on 114 adults showed that 8 weeks after starting SP, the intervention group reported decreased loneliness with a large effect size^15^. Retention of those allocated to SP was also high (79.4%) ^15^. However, applications of youth SP for loneliness using more robust methods are yet to be undertaken.

Referral routes into SP have traditionally been through primary care, yet research suggests that younger populations are more likely to access SP via non-medical routes^17^. Schools offer potential as a non-medical route to refer YP into SP as they represent a universal point of access to wellbeing support for many YP^18^. To our knowledge, SP has not been formally implemented or evaluated in schools. A recent report by the National Academy for Social Prescribing highlighted that educational stakeholders have been underrepresented in SP pathway development and that to be successful, work needs to be undertaken to ensure adequate information sharing between agencies, such as schools and LWs^19^.

Given the high prevalence of loneliness among YP and the need for tailored, accessible interventions, the INcreasing Adolescent social and Community supporT (INACT) study piloted a novel school-based SP pathway for pupils reporting elevated loneliness. The aims were to (i) assess feasibility, acceptability and appropriateness of the pathway and trial procedures; (ii) test the proposed measurement framework; and (iii) generate parameters to inform the design and sample size of a future definitive trial.

## Materials and methods

### Design

The research design consisted of a two-group (SP vs. signposting) parallel randomised design, with YP as the unit of randomisation. Ethical approval was granted from UCL Research Ethics Committee (6735/017). INACT was prospectively registered on ClinicalTrials.gov (NCT06656663) on 18/10/2024 and a pilot protocol publication outlines the study design and methods^20^. The study broadly followed the registered protocol, although higher-than-anticipated loneliness prevalence, together with operational constraints related to link worker capacity, led to a pragmatic deviation from equal allocation during implementation. This paper adheres to the CONSORT extension for randomised pilot and feasibility trials^21^ (see supplementary material: S0). A youth advisory group co-informed the study design, outcome measure selection, and interpretation of findings, ensuring relevance and acceptability to YP.

### Sample size

As this was a pilot trial, the primary objective was to assess feasibility and estimate parameters for a future definitive trial rather than to detect statistically significant differences in outcome measures. A target sample of 78 participants was pragmatically determined based on anticipated school size, loneliness prevalence, and LW capacity.

### Setting

INACT was conducted in 12 mainstream schools (six primary and six secondary) across three urban areas in England (London, Leeds, and Manchester). In each area, four schools participated (two primary and two secondary). These cities were selected to reflect urban contexts characterised by socioeconomic diversity and higher levels of deprivation, factors associated with increased risk of loneliness^4^.

### Participants

Participants were pupils in either Years 4 or 5 (primary school) or Years 7 or 8 (secondary school) who were reporting elevated loneliness; they were on average 10.93 years of age. A threshold of >6 on the Good Childhood Index loneliness was used to indicate elevated loneliness. YP deemed by school staff to have a severe learning disability were excluded from the study due to potential difficulties with questionnaire completion.

### Intervention and counterfactual

The SP intervention used in INACT was ‘YES’: Youth Engagement in SP. It consisted of approximately six sessions delivered over an eight-week period, which took place online, via telephone, or in person depending on each YP’s preference. During sessions, LWs explored ‘what matters’ to YP and used this information, alongside their knowledge of local community assets, to co-develop a personalised plan to connect YP to sources of support within their local area. With YP agreement, LWs facilitated engagement with identified activities. This included introducing YP to relevant services and, where appropriate, accompanying them to an initial session. Each LW held an “enabling grant” (a personalised discretionary budget of up to £40 per YP) to address practical barriers to engagement, such as equipment, travel, or activity costs. All LWs received training and ongoing supervision from an experienced health professional, such as a clinical psychologist, and a senior LW.

YP in the control group received signposting to social and community activities in their local area. School pastoral staff provided YP and their parents or guardians with a leaflet detailing information on the same local sources of support identified through LW asset mapping.

### Pupil measures

The primary outcome measure for INACT was the three loneliness questions from the Good Childhood Index^22^. Each question is scored on a three-point Likert scale ranging from 1 (“hardly ever or never”) to 3 (“often”), with higher total scores indicating greater loneliness. Loneliness was assessed at baseline, three- and six-months, with three-month follow-up specified as the primary timepoint examined here. A threshold of >6 was used to indicate elevated loneliness, corresponding to a conservative threshold above occasional loneliness. Other measures were selected with YP and subject expert input. Further details on constructs and measures are provided in the Supplementary Material (S1).

### Implementation and process monitoring

Fidelity to the YES SP model was supported through standardised training, supervision, and documentation of session content. Routine monitoring of the number of sessions delivered, delivery mode, and referral activity was undertaken by LWs. For school staff and LWs, feasibility, acceptability and appropriateness of SP was assessed with the Feasibility of Implementation Measure, Acceptability of Intervention Measure and Appropriateness of Intervention Measure^23^. Each questionnaire consisted of four questions, each on a five-point Likert scale ranging from 1 ‘Completely Disagree’ to 5 ‘Completely Agree’. Higher scores indicate greater feasibility, acceptability, and appropriateness. Because these measures have not been validated for use with YP, acceptability of SP was assessed by the proportion of YP who took up the offer, whilst acceptability of the measures was assessed by the proportion of YP completing study measures at baseline and three months.

### Qualitative data

Qualitative interviews with YP, LWs, and school staff were conducted online via Microsoft Teams to explore the acceptability of SP, perceived barriers and facilitators, and perceptions of the study design. YP were purposively sampled based on age, gender, ethnicity, site, loneliness score, and level of engagement with SP. LWs and school staff were sampled based on location. Interviews with YP took place at least three-months after SP initiation, while interviews with LWs and staff occurred at least 4 months after initiation. Interview guides for LWs and school staff focused on delivery, perceived impact, and intention to adopt or continue SP or signposting. Guides for YP explored acceptability, feasibility, and suitability of SP, as well as engagement with community support following referral. Interview guides are available in the Supplementary Materials (see S2). Overall, five LWs involved in the pilot were invited to interview, of whom three participated. School staff involved in delivery were also invited, but no interviews were completed despite follow-up contact. Five YP participated in interviews. Characteristics of YP and LWs participating in interviews are provided in the Supplementary Materials (S3 and S4).

### Harm monitoring

Adverse events were defined *a priori* as any safeguarding concern, deterioration in wellbeing requiring school or clinical escalation, or any serious untoward event occurring during participation. Schools and LWs were instructed to report any such events to the research team and local safeguarding leads, with any reported events reviewed by the Chief Investigator for escalation. No adverse events were reported during the pilot.

### Procedure

Schools selected whole classes of pupils to participate in INACT (one class from Years 4 and 5 in primary schools and, two classes from Years 7 and 8 in secondary schools). Opt-out letters were sent to parents/guardians giving them two weeks to declare to the study team that they did not want their child to participate. Participant recruitment and follow-up data collection took place between November 2024 and June 2025. For pupils who were not opted out, schools then provided the research team with the name, gender, class name and unique pupil number (UPN) of each pupil. Pupils were then able to complete the baseline survey, if they assented to do so, at an appropriate time as deemed by the school, such as in their Personal, Social, Health and Economic lessons. Following completion of baseline surveys, the research team performed screening using the Good Childhood Index^22^. YP meeting the predetermined threshold for loneliness (scoring >6) were then randomly allocated by the study statistician to either receive SP or signposting. Randomisation was conducted by a member of the research team [removed for blinding] using a computer-generated random allocation sequence. Due to operational constraints, including LW capacity, allocation was not strictly 1:1 in practice. The number assigned to the SP arm was capped at 41 to ensure feasible delivery of the intervention and to pilot the referral and delivery process, while still allowing estimation of comparative parameters (e.g. effect sizes and intraclass correlation coefficients) to inform a future definitive trial. All remaining eligible participants were allocated to the signposting group.

For follow-up data collection, a matched subset of participants in the signposting group was randomly selected (1:1 relative to the SP group). This sub-sampling occurred after random allocation and was undertaken for follow-up efficiency rather than outcome-based selection. All participants allocated to SP were included in follow-up.

Allocation was concealed from school staff and LWs until the point of assignment. The allocation list was held securely by the research team and communicated only after all pupils had completed baseline surveys. School staff then contacted parents/guardians and pupils to inform them of their allocation. Those allocated to SP were provided with an outline of what it entailed using a standardised script and those that were interested in receiving SP had their contact details passed onto the LW. Those allocated to signposting were provided with a leaflet by school staff which had been co-developed with a LW and detailed sources of support in the community. All YP were followed up at three- and six-months post allocation.

Due to the nature of the intervention, blinding of participants was not possible, as allocation determined whether YP received active SP sessions or signposting only. Data analysts were not blinded to group assignment during primary modelling, as this was a pilot feasibility study focused primarily on implementation and estimation of preliminary effects.

For qualitative data, interviews were conducted online via Microsoft Teams. With participants’ assent/consent, interviews were audio-recorded and transcribed verbatim. Identifiable information was removed prior to analysis.

### Analysis

To assess the feasibility, acceptability and suitability of the SP pathway, descriptive statistics were calculated for stakeholders’ questionnaire responses and uptake of the SP offer, alongside qualitative interview analysis. Descriptive quantitative data was analysed with Stata^24^, whilst qualitative data were managed in NVivo^25^ and analysed using framework analysis^26^. Transcripts were coded by the lead researcher [removed for blinding], with a subset double coded by a second researcher [removed for blinding]. A coding framework was developed iteratively, combining deductive and inductive approaches, and applied across all transcripts. Analytical matrices were developed and reviewed by the Youth Advisory Group team to support interpretation and ensure consistency.

To assess whether the preliminary power calculation assumptions were realistic (see Supplementary Material: S5), data from point estimates of the within-person intraclass correlation coefficients (ICCs) were calculated, as well as the effect size of the SP intervention. To estimate preliminary intervention effects, Bayesian growth curve modelling was used to examine changes in loneliness across three time points, with random intercept and slope. Time was used as a discrete variable to allow non-linear growth trajectories with group (SP vs signposting) included as a fixed effect. Missing outcome data were handled within the Bayesian mixed-effects framework using full information maximum likelihood estimation under a missing-at-random assumption. No imputation was undertaken. Sensitivity analyses were conducted using an alternative measure of peer loneliness.

No interim efficacy analyses were conducted. Progression to a definitive trial was guided by predefined feasibility stop–go criteria (see Supplementary Material: S6).

## Results

### Participant flow and recruitment

Figure 1 shows the recruitment and retention of schools and pupils in INACT. Of the 12 schools recruited, one primary school dropped out due to the perceived suitability of the validated measures selected by the research team and youth advisory group. The remaining schools sent opt-out letters to parents/guardians of 920 potential pupils and 19 (2%) opted their YP out. When asked to assent to the study prior to baseline completion, 229 (25%) pupils decided not to participate.

**Figure 1.**
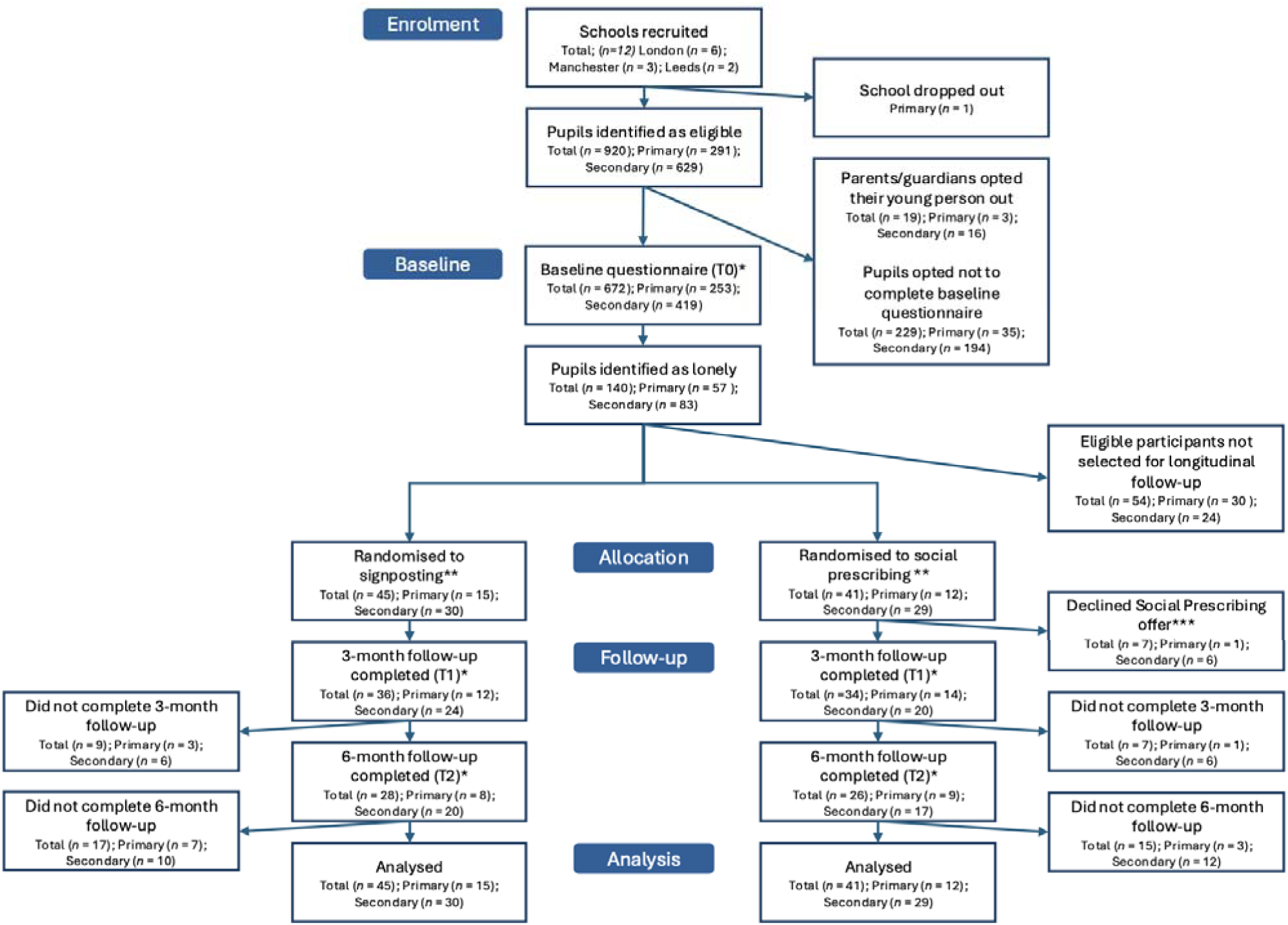
CONSORT flow diagram of participant recruitment, allocation, follow-up and analysis.

Of the 672 participants who completed baseline measures, 140 (20.8%) scored greater than six on the Good Childhood Index^19^. In total, 86 participants were enrolled in the pilot, including 45 in the signposting group and 41 in the SP group. At three months, 70 (81%) participants completed follow-up, whilst at six-months, 54 participants (62%) completed follow-up. Reasons for attrition included pupil absence on follow-up day, withdrawal of assent, and school-level scheduling constraints. No withdrawals were due to adverse events.

### Sample characteristics

The socio-demographic characteristics of the trial participants are described in Table 1. The mean age of participants was 10.87 (standard deviation [SD]=1.85) with just over half (58%) identifying as a girl and just over half (52%) identifying as White. In total, fourteen percent identified as having a disability and 54% lived with both parents. Just over two-thirds (69%) of pupils were from secondary schools. The signposting and SP groups had broadly similar characteristics.

**Table 1.** Sample characteristics.

|  | Total<br>(n=86) | Signposting<br>(n=45) |  |  | SP<br>(n=41) |  |
| --- | --- | --- | --- | --- | --- | --- |
|  | Freq | Mean<br>(SD)/% | Freq | Mean<br>(SD)/% | Freq | Mean<br>(SD)/% |
| Age | 86 | 10.87<br>(1.85) | 45 | 10.82<br>(1.97) | 34 | 10.93<br>(1.72) |
| Gender<sup>†</sup>:<br>boy | 33 | 41.3% | 18 | 40.9% | 15 | 41.7% |
| Gender<sup>†</sup>:<br>girl | 47 | 58.8% | 26 | 59.1% | 21 | 58.3% |
| Ethnicity:<br>white | 36 | 52.2% | 17 | 50.0% | 19 | 54.3% |
| Ethnicity:<br>other | 33 | 47.8% | 17 | 50.0% | 16 | 45.7% |
| Ill-<br>ness/disabi<br>lity: yes | 12 | 14.0% | 7 | 15.6% | 5 | 12.2% |
| Ill-<br>ness/disabi<br>lity: no | 40 | 46.5% | 21 | 46.7% | 19 | 46.3% |
| Ill-<br>ness/disabi<br>lity: not<br>sure | 34 | 39.5% | 17 | 37.8% | 17 | 41.5% |
| Living<br>with: both<br>parents | 46 | 53.5% | 23 | 51.1% | 23 | 56.1% |
| Living<br>with: one<br>parent | 27 | 31.4% | 14 | 31.1% | 13 | 31.7% |
| Living<br>with: other | 13 | 15.1% | 8 | 17.8% | 5 | 12.2% |
| School:<br>primary | 27 | 31.4% | 15 | 33.3% | 12 | 29.3% |
| School:<br>secondary | 59 | 68.6% | 30 | 66.7% | 29 | 70.7% |

Notes: Pupils self-identified as non-binary or other gender were coded as missing due to small numbers. Fre- quencies may not sum to group totals due to item-level missing data. Percentages are based on available data.

Participants’ scores on primary and secondary outcome measures at baseline, three-and six- months are outlined in Table 2. At baseline, participants’ mean loneliness score on the Good Childhood Index^19^ was 7.60 (SD=0.69) in the signposting group and 7.71 (SD=0.87) in the SP group. At baseline, participants reported elevated perceived stress (mean scores around 9 on a 0–16 scale). Baseline values for all primary and secondary outcome measures were broadly similar between the signposting and SP groups.

**Table 2.** Participant outcomes: Baseline, 3-, and 6-months.

| Outcomes | Baseline Mean (SD) |  | 3 months Mean (SD) |  | 6 months Mean (SD) |  |
| --- | --- | --- | --- | --- | --- | --- |
|  | Signposting | SP | Signposting | SP | Signposting | SP |
| Loneliness (Good Childhood Index)<sup>22</sup> | 7.60<br>.69 | 7.71<br>.87 | 5.74<br>1.58 | 5.28<br>1.86 | 5.44<br>1.52 | 5.87 2.00 |
| Peer loneliness Loneliness and Aloneness Scale for Children and Adolescents (LACA)<sup>27</sup> | 32.42<br>6.85 | 33.02<br>6.68 | 26.76<br>6.78 | 25.36<br>9.06 | 26.10<br>7.47 | 26.17 8.21 |
| Stress (Perceived Stress Scale)<sup>28</sup> | 9.33<br>2.17 | 8.93<br>3.06 | 8.70<br>1.95 | 8.00<br>2.64 | 9.00<br>2.00 | 8.05<br>3.12 |
| Wellbeing (Kidscreen)<sup>29</sup> | 18.64<br>6.46 | 17.66<br>6.14 | 19.33<br>5.61 | 20.33<br>6.30 | 20.93<br>5.17 | 20.53 6.03 |
| Emotional difficulties (Me & MyFeelings)<sup>30</sup> | 10.98<br>3.79 | 10.69<br>4.66 | 7.67<br>3.58 | 7.83<br>3.84 | 7.34<br>4.07 | 7.77<br>4.50 |

### Intervention engagement and fidelity

For the 41 YP allocated to SP, 34 (83%) attended at least one SP session with a LW. Across participants who started SP, the mean number of LW sessions attended was 5.09 (SD=1.42) and 90% of sessions took place in person. Of the 34 participants, 16 (47%) were referred onto a community activity and the personalised budget was used with 26 of the 34 participants (76%). No adverse events were reported. No major deviations from the intended delivery model were identified.

### Feasibility, acceptability, and appropriateness (quantitative)

Data from the quantitative measures on intervention feasibility, acceptability and appropriateness^23^ from the perspectives of school staff and LWs are shown in Table 3. Overall, both school staff and LWs felt that SP for lonely pupils was feasible, acceptable, and appropriate, with LWs agreeing more strongly with each area when compared to school staff.

**Table 3.** LW and school staff views on SP.

|  | LWs (n=3)<br>Mean (SD) | School Staff (n=8)<br>Mean (SD) |
| --- | --- | --- |
| Feasibility (1-5)* | 4.75 (0.45) | 4.06 (0.61) |
| Acceptability (1-5) | 4.67 (0.49) | 4.09 (0.64) |
| Appropriateness (1-5) | 4.58 (0.51) | 4.13 (0.63) |

\*Higher scores equal more positive ratings

### Effect estimates and preliminary efficacy

The within person ICC for the Good Childhood Index^22^ was 0.13. All participants in the longitudinal analytic sample were analysed according to their allocated group, irrespective of intervention uptake, consistent with an intention-to-treat principle. Figure 2 shows the predicted trajectories of loneliness across three time points by intervention group (see also Supplementary Material S7). Loneliness decreased between baseline and three-month follow-up in both groups. Specifically, loneliness decreased by 1.83 points (95% CI: -2.45 to -1.22) in the signposting group and by 2.12 points (95% CI: -2.79 to -1.46) in the SP group (Figure 2a). Although the point estimate suggested a greater reduction in loneliness in the SP group at three-months, the between-group difference in change was imprecisely estimated and the confidence interval included no clear difference between groups (coef = -0.57, 95% CI: -1.46 to 0.33). This corresponded to a standardised effect size of 0.32, with a wide confidence interval that included no effect (95% CI: -0.81 to 0.17) (Figure 2b). However, findings at subsequent follow-up suggested that any potential impact of SP on loneliness may not have been maintained. Observed ICC and effect size estimates were broadly consistent with assumptions used for preliminary power calculations and provide parameters to inform a future definitive trial. Analyses using an alternative measure of peer loneliness showed improvements over time in both groups, with no clear evidence of between-group differences (Supplementary Material S8). Sensitivity analyses excluding participants who did not receive SP, and additionally excluding those who started later than planned and still had ongoing sessions at three-month follow-up, produced materially similar estimates and did not alter substantive conclusions (Supplementary Material S9).

**Figure 2.**
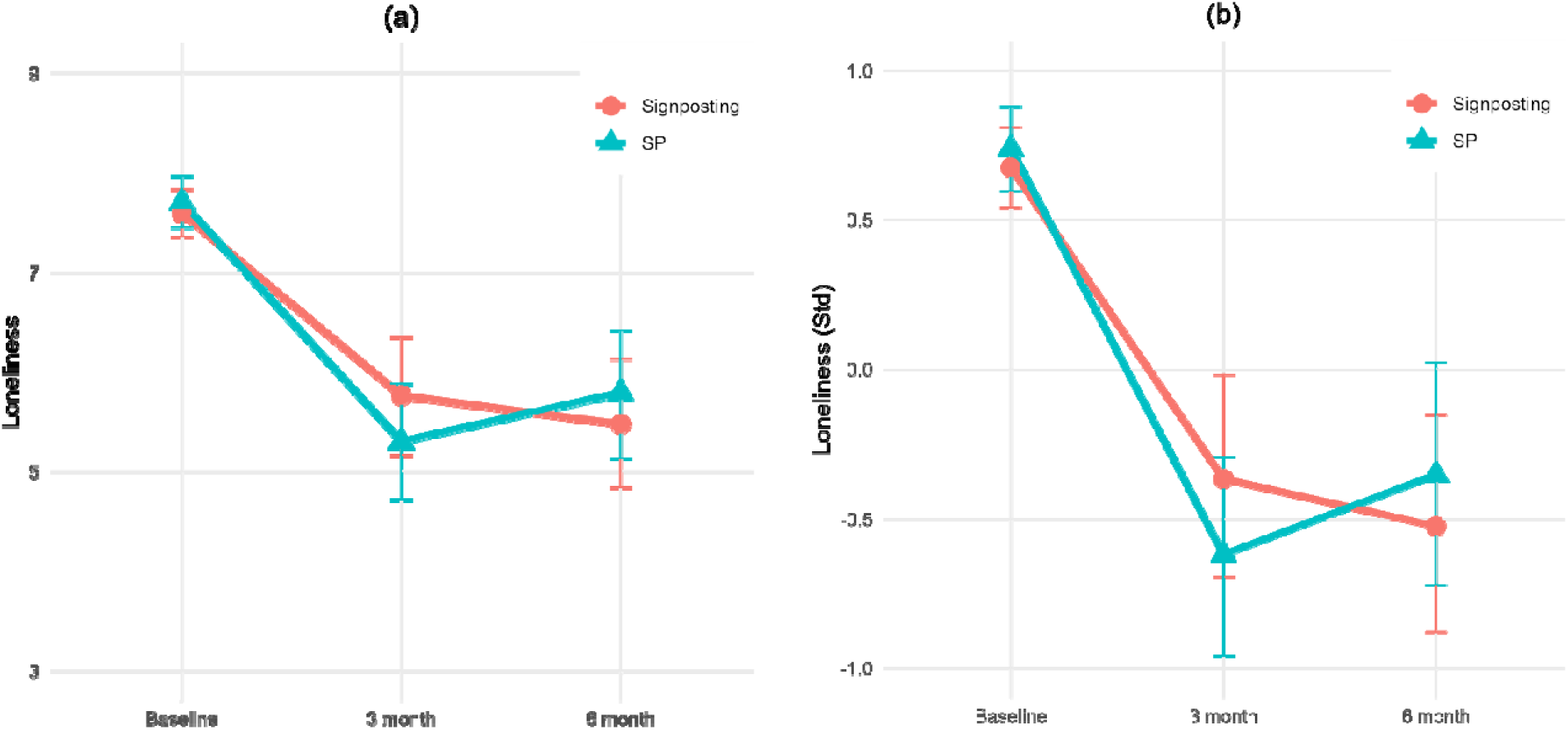
Predicted loneliness trajectories from Bayesian growth curve model.

### Qualitative findings: feasibility and acceptability of the SP pathway

Qualitative findings draw on interviews with three LWs and five YP; no school staff interviews were completed. Link worker interviews lasted between 25 and 64 minutes (mean 41.8 minutes). Five young people also participated in interviews, which lasted between 7 and 24 minutes (mean 16.0 minutes) LWs described the school-based SP pathway as broadly feasible when supported by three interrelated conditions: parental engagement, school facilitation, and flexibility in delivery. Parent and family engagement was often described as enabling uptake and continued participation:

*“They engage well in conversations and also parent engagement was very good as well…A lot of parents are on board with what we’re trying to achieve” (LW_001)*

Schools were also viewed as important implementation contexts. Access to school-based space reduced travel burden, supported continuity, and enabled practical delivery:

*“I was able to offer the sessions in school…I was in one place…with my resources…not having to travel” (LW_002)*

Together, these accounts suggest feasibility appeared to depend not simply on the intervention itself, but on the relational and organisational conditions surrounding delivery.

Feasibility was also shaped by contextual barriers, particularly family capacity and practical constraints affecting engagement. LWs suggested younger pupils often depended on parental support to attend sessions or engage with referred activities, which could be more difficult in some family circumstances:

*“Kids with lots of siblings…single parent families…had less capacity” (LW_003)*

However, LWs also described adapting to these constraints through flexible arrangements, including enabling grants, sensitivity to family context, and use of school-based delivery to reduce practical barriers. These accounts suggest barriers were often dynamic, interacting with adaptive responses from LWs. For some YP, familiar school settings also appeared to support comfort and engagement.

*“Most of the meetings were at my school… it felt I knew where I was… it was more convenient.” (YP_001)*

YP generally found SP acceptable, particularly valuing relational aspects of support and the personalised focus on their interests. Several YP appeared to value continuity in meeting with the same LW over repeated sessions, which seemed to support trust and engagement over time:

*“At first… it got a bit hard to socialise with him but then it got easier… we kept talking and it just sort of clicked.” (YP_004)*

Participants often described sessions as enjoyable and highlighted feeling listened to and understood:

*“[LW] was really fun…I liked how [they] asked me what my hobbies were…we did that sort of every session” (YP_001)*

Interactive activities, including games and strengths-focused exercises, were often viewed positively and sometimes appeared to support self-recognition of interests and capabilities.

Acceptability was not uniform. Some YP disliked particular activities, especially writing-based exercises, while others suggested sessions could be too long or that meeting out-side school might have been preferable:

*“The writing, yeah, I didn’t really like it” (YP_005)*

*“I didn’t enjoy how long it was” (YP_002)*

These accounts suggested preferences for flexibility in session format, activity modality, and meeting location.

### Acceptability of study procedures

LWs provided limited feedback on the study procedures themselves. One described research processes as generally smooth, though noted some duplication in documentation requirements.

*“it was more duplication of my work because I documented the interventions anyway…I obviously had to do it again to provide evidence for the [INACT] project” (LW_001)*

YP offered few concerns regarding participation in the study, and suggestions focused more on refining intervention delivery, described above, than the research procedures. Overall, findings suggested study procedures were acceptable. Prespecified progression criteria were met overall, supporting progression to a definitive trial, albeit with modifications to improve follow-up retention.

## Discussion

This pilot randomised controlled trial provides the first trial evidence that a SP pathway delivered through schools for YP experiencing loneliness is feasible, appropriate, and acceptable, with preliminary signals of short-term benefit. Embedding SP within mainstream educational settings enabled successful identification, engagement, and delivery of personalised support to lonely pupils, addressing a key limitation of existing pathways that are largely accessed through primary care^17^. Although reductions in loneliness were observed in both intervention conditions, participants receiving SP showed numerically greater reductions at three months, suggesting potential added value from relational, facilitated support beyond signposting alone. However, effects were modest and not clearly sustained at six-months, reinforcing that the primary contribution of this study is to establish implementation feasibility and generate effect estimates to inform a fully powered definitive trial.

Screening for loneliness within schools was achievable, with approximately one in five screened pupils meeting eligibility criteria. This prevalence was higher than national estimates (11.3%) but likely reflects recruitment in urban areas with higher deprivation, where loneliness risk may be elevated^4^. Uptake of SP was high, with 83% initiating the pathway, broadly comparable to adult SP pathways for loneliness^15^. Three-month follow-up completion (81%) supports the short-term feasibility of the assessment approach within a school-based trial context. However, attrition at six-months (62%) suggests that enhanced retention strategies will be important in a definitive trial.

Implementation measures indicated that both LWs and school staff perceived the pathway as feasible, acceptable and appropriate, with particularly strong endorsement from LWs. This may reflect that LWs already operate across community assets as part of their role^17,31^, whereas for school staff this involved some additional administrative adjustments, such as room booking. Nonetheless, both groups positively endorsed these measures. Qualitative findings added explanatory depth, highlighting the importance of relational continuity, personalised activity matching, and flexibility in delivery. Schools appeared to function not simply as referral sites, but as supportive settings for engagement, including for YP hesitant about unfamiliar community venues. Together, these findings support the potential of schools as accessible infrastructure for delivering non-clinical wellbeing interventions^18^.

Implementation challenges were also identified. Younger pupils often depended on parental capacity to engage with community activities, and complex family circumstances sometimes constrained participation. Practical organisational challenges, including duplication of documentation, were also noted. These issues have implications for scalability and suggest that clearer communication pathways, streamlined processes, and integrated monitoring systems may be important in a future trial. In addition, improvements observed in both groups may partly reflect regression to the mean following eligibility screening based on elevated loneliness, which should be considered in interpreting preliminary outcome signals.

Both SP and signposting groups demonstrated reductions in loneliness over three months. Although reductions were somewhat greater in the SP group, between-group differences were imprecisely estimated, as expected in a pilot study. The observed standardised effect size (0.32) is broadly consistent with moderate effects reported in meta-analyses of youth loneliness interventions^9,10^. It is notable that signposting alone was also associated with reductions in loneliness. This may reflect increased awareness of available activities, behavioural activation prompted by screening and referral, or regression to the mean. The potential added value of SP may lie in its relational and facilitative components, including co-produced planning, accompaniment to initial sessions, and ongoing motivational support^31,32^. The attenuation of effects by six-months may suggest a need for longer engagement, booster input or closer integration within school wellbeing systems.

To our knowledge, this is the first pilot randomised trial evaluating a structured SP pathway for loneliness delivered via schools. Previous SP evaluations have predominantly focused on adults and have frequently lacked comparison groups^14^. By embedding SP within an educational context and employing a parallel randomised design, this study contributes preliminary evidence regarding both implementation feasibility and potential effectiveness in a youth population. The findings also respond to calls for more tailored approaches to youth loneliness^9,10^. Unlike universal classroom-based programmes, which may offer limited tailoring to differing loneliness needs^9^, SP offers individualised support shaped around YP’s interests, preferences and circumstances. This flexibility may be particularly important given the socially patterned nature and heterogeneity of YP’s experiences of loneliness^4^.

The study also supported the provisional measurement framework, including acceptable use of proposed outcomes, estimation of ICC and effect size parameters, and identification of refinements needed for younger pupils. Strengths of this study include its multi-site design across three urban areas and inclusion of both primary and secondary school pupils. The integration of quantitative implementation measures with qualitative interviews enabled a more comprehensive assessment of feasibility and acceptability. However, several limitations should be acknowledged. LW capacity constrained intervention allocation, follow-up was relatively short, and school staff did not participate in qualitative interviews, limiting insight into organisational perspectives. Outcomes relied on self-report, and staff reported back to the research team that some younger pupils required assistance completing questionnaires, suggesting that measurement burden and developmental suitability warrant careful consideration in a future trial.

Overall, these findings suggest that screening for loneliness and delivering tailored SP within schools is feasible and acceptable, with early signals of benefit. A fully powered multi-site trial with economic evaluation is now warranted to determine effectiveness, test mechanisms of change and assess longer-term sustainability. Given schools’ near-universal reach, embedding relational, community-connecting interventions within educational settings may offer a scalable complement to primary care-based SP pathways.

## Statements and Declarations

### Ethical considerations

Ethical approval for this study was granted by the University College London Research Ethics Committee (Reference: 6735/017). The study was conducted in accordance with relevant ethical guidelines and regulations.

### Consent to participate

Parental opt-out procedures and young person assent were used for pupil participants in accordance with ethics approval. Written informed consent was obtained from adult interview participants.

### Consent for publication

Not applicable.

## Data availability

An anonymised quantitative dataset underlying the findings is being pre-pared for controlled sharing, and arrangements for an appropriate repository are currently being discussed with the study funders. Pending completion of those arrangements, de-identified quantitative data may be available from the corresponding author on reasonable request and subject to appropriate approvals. Qualitative interview data are not publicly available due to confidentiality and participant identifiability considerations.

## Data Availability

An anonymised quantitative dataset underlying the findings is being prepared for controlled sharing, and arrangements for an appropriate repository are currently being discussed with the study funders. Pending completion of those arrangements, de-identified quantitative data may be available from the corresponding author on reasonable request and subject to appropriate approvals. Qualitative interview data are not publicly available due to confidentiality and participant identifiability considerations.

## Acknowledgements

We would like to thank the SPYN Youth Advisory Group for their valuable input and support throughout this study.

## References

1. World Health Organisation. From loneliness to social connection Charting a path to healthier societies Report of the WHO Commission on Social Connection. Geneva, https://www.who.int/publications/i/item/978240112360 (2025, accessed 12 February 2026).

2. Nicolaisen M, Thorsen K. Who are Lonely? Loneliness in Different Age Groups (18–81 Years Old), Using Two Measures of Loneliness. The International Journal of Aging and Human Development 2014; 78: 229–257.

3. Pyle E, Evans D. Loneliness - What characteristics and circumstances are associated with feeling lonely? Analysis of characteristics and circumstances associated with loneliness in England using the Community Life Survey, 2016 to 2017. London, https://www.ons.gov.uk/peoplepopulationandcommunity/wellbeing/articles/lonelinesswhatcharacteristicsandcircumstancesareassociatedwithfeelinglonely/2018-04-10?utm_source=chatgpt.com (2018, accessed 12 February 2026).

4. Office for National Statistics. Children’s and young people’s experiences of loneliness: 2018. London, 2018.

5. Qualter P, Brown SL, Munn P, et al. Childhood loneliness as a predictor of adolescent depressive symptoms: an 8-year longitudinal study. Eur Child Adolesc Psychiatry 2010; 19: 493–501.

6. Caspi A, Harrington H, Moffitt TE, et al. Socially Isolated Children 20 Years Later. Arch Pediatr Adolesc Med 2006; 160: 805.

7. Xerxa Y, Rescorla LA, Shanahan L, et al. Childhood loneliness as a specific risk factor for adult psychiatric disorders. Psychol Med 2023; 53: 227–235.

8. Hawkley LC, Cacioppo JT. Loneliness Matters: A Theoretical and Empirical Review of Consequences and Mechanisms. Annals of Behavioral Medicine 2010; 40: 218–227.

9. Eccles AM, Qualter P. Review: Alleviating loneliness in young people – a metaLanalysis of interventions. Child Adolesc Ment Health 2021; 26: 17–33.

10. Burke L, Christiansen J, Lasgaard M, et al. Interventions to reduce loneliness in children and adolescents (4–18 years): A systematic review and meta-analysis with narrative synthesis of study-level characteristics. J Affect Disord 2026; 403: 121445.

11. National Academy for Social Prescribing. What is Social Prescribing? National Academy for Social Prescribing, https://socialprescribingacademy.org.uk/what-is-social-prescrib-ing/#:~:text=Social%20prescribing%20connects%20people%20to,on%20what%20works%20for%20them.https://www.england.nhs.uk/personalisedcare/workforce-and-training/social-prescribing-link-work-ers/#:~:text=Social%20prescribing%20link%20workers%20connect,housing%2C%20financial%20and%20welfare%20advice. (2021, accessed 13 August 2023).

12. Jones D, Jopling K, Kharicha k. Loneliness beyond Covid-19: Learning Lessons of the Pandemic for a Less Lonely Future. London, https://www.campaigntoendloneliness.org/wp-content/uploads/Loneliness-beyond-Covid-19-July-2021.pdf (2021, accessed 13 August 2023).

13. Hayes D, Jarvis-Beesley P, Mitchell D, et al. The impact of social prescribing on children and young people’s mental health and wellbeing’. . London, https://socialprescribingacademy.org.uk/media/lrif2emh/evidence-review-the-impact-of-social-prescribing-on-children-and-young-peoples-health-and-wellbeing.pdf (2023, accessed 21 June 2023).

14. Bickerdike L, Booth A, Wilson PM, et al. Social prescribing: less rhetoric and more reality. A systematic review of the evidence. BMJ Open 2017; e013384.

15. Dingle G, Sharman LS, Hayes S, et al. A controlled evaluation of social prescribing on loneliness for adults in Queensland: 8-week outcomes. Front Psychol 2024; 15: 1359855.

16. Foster A, Thompson J, Holding E, et al. Impact of social prescribing to address loneliness: A mixed methods evaluation of a national social prescribing programme. Health Soc Care Community 2021; 29: 1439–1449.

17. Bu F, Hayes D, Burton A, et al. Equal, equitable or exacerbating inequalities? Patterns and predictors of social prescribing referrals in 160,128 UK patients. The British Journal of Psychiatry 2025; 2: 91–99.

18. Aviles AM, Anderson TR, Davila ER. Child and adolescent socialLemotional development within the context of school. Child Adolesc Ment Health 2006; 11: 32–39.

19. Polley P, Hayes D, Husk K. National Survey of Children and Young People’s Social Prescribing in England 2022 snapshot.

20. Hayes D, Burton A, Bu F, et al. INcreasing Adolescent social and Community supporT (INACT): Pilot study protocol. PLoS One 2025; 20: e0317823.

21. Eldridge SM, Chan CL, Campbell MJ, et al. CONSORT 2010 statement: extension to randomised pilot and feasibility trials. BMJ 2016; i5239.

22. The Children’s Society. Loneliness in childhood: Exploring loneliness and well-being among 10–17 year olds. London, 2019.

23. Weiner BJ, Lewis CC, Stanick C, et al. Psychometric assessment of three newly developed implementation outcome measures. Implementation Science 2017; 12: 108.

24. StataCorp. Stata Statistical Software: Release 17.

25. QSR International Pty Ltd. NVivo.

26. Parkinson S, Eatough V, Holmes J, et al. Framework analysis: a worked example of a study exploring young people’s experiences of depression. Qual Res Psychol 2016; 13: 109–129.

27. Goossens L, Maes M. Loneliness and Aloneness Scale for Children and Adolescents (LACA). In: Encyclopedia of Personality and Individual Differences. Cham: Springer International Publishing, 2017, pp. 1–5.

28. Demkowicz O, Panayiotou M, Ashworth E, et al. The Factor Structure of the 4-Item Perceived Stress Scale in English Adolescents. European Journal of Psychological Assessment 2020; 36: 913–917.

29. Ravens-Sieberer U, Gosch A, Rajmil L, et al. KIDSCREEN-52 quality-of-life measure for children and adolescents. Expert Rev Pharmacoecon Outcomes Res 2005; 5: 353–364.

30. Deighton, J., Tymms, P., Vostanis, P., Belsky, J., Fonagy, P., Brown, A., Martin, A., Patalay, P. & Wolpert W. The Development of a School-Based Measure of Child Mental Health. J Psychoeduc Assess 2013; 31: 247–257.

31. NHS England. Social Prescribing Link Workers. NHS England, https://www.england.nhs.uk/personalisedcare/workforce-and-training/social-prescribing-link-work-ers/#:~:text=Social%20prescribing%20link%20workers%20connect,housing%2C%20financial%20and%20welfare%20advice. (2021, accessed 13 August 2023).

32. NHS. Social Prescribing Link Worker Competency Framework. 2022.

