## Supplementary Materials for "School-based social prescribing to address youth loneliness: a pilot randomised controlled trial"

S0: CONSORT 2010 checklist of information to include when reporting a pilot or feasibility trial

S1: Secondary outcomes, moderators, and mechanism measures

S2: Interview Guides

S3: Characteristics LWs participating in interviews

S4: Characteristics of YP participating in interviews

S5: Preliminary power calculation

S6: Stop-Go criteria

S7: Main loneliness model

S8: Peer loneliness model

S9: Sensitivity analyses

**S0: CONSORT 2010 checklist of information to include when reporting a pilot or feasibility trial**

| Section/Topic | Item No | Checklist item | Reported on page No |
| --- | --- | --- | --- |
| Title and abstract | | | |
|  | 1a | Identification as a pilot or feasibility randomised trial in the title | Title page |
|  | 1b | Structured summary of pilot trial design, methods, results, and conclusions (for specific guidance see CONSORT abstract extension for pilot trials) | 1 |
| Introduction | | | |
| Background and objectives | 2a | Scientific background and explanation of rationale for future definitive trial, and reasons for randomised pilot trial | 2-3 |
|  | 2b | Specific objectives or research questions for pilot trial | 3 |
| Methods | | | |
| Trial design | 3a | Description of pilot trial design (such as parallel, factorial) including allocation ratio | 4 |
|  | 3b | Important changes to methods after pilot trial commencement (such as eligibility criteria), with reasons | 5 |
| Participants | 4a | Eligibility criteria for participants | 4-5 |
|  | 4b | Settings and locations where the data were collected | 4 |
|  | 4c | How participants were identified and consented | 6-7 |
| Interventions | 5 | The interventions for each group with sufficient details to allow replication, including how and when they were actually administered | 5-6 |
| Outcomes | 6a | Completely defined prespecified assessments or measurements to address each pilot trial objective specified in 2b, including how and when they were assessed | 6-7; S1 |
|  | 6b | Any changes to pilot trial assessments or measurements after the pilot trial commenced, with reasons | N/A |
|  | 6c | If applicable, prespecified criteria used to judge whether, or how, to proceed with future definitive trial | S6 |
| Sample size | 7a | Rationale for numbers in the pilot trial | 4 |
|  | 7b | When applicable, explanation of any interim analyses and stopping guidelines | 8 |
| Randomisation: |  |  |  |
| Sequence  generation | 8a | Method used to generate the random allocation sequence | 7 |
|  | 8b | Type of randomisation(s); details of any restriction (such as blocking and block size) | 7 |
| Allocation  concealment  mechanism | 9 | Mechanism used to implement the random allocation sequence (such as sequentially numbered containers), describing any steps taken to conceal the sequence until interventions were assigned | 7 |
| Implementation | 10 | Who generated the random allocation sequence, who enrolled participants, and who assigned participants to interventions | 7 |
| Blinding | 11a | If done, who was blinded after assignment to interventions (for example, participants, care providers, those assessing outcomes) and how | 7-8 |
|  | 11b | If relevant, description of the similarity of interventions | N/A |
| Statistical methods | 12 | Methods used to address each pilot trial objective whether qualitative or quantitative | 8 |
| Results | | | |
| Participant flow (a diagram is strongly recommended) | 13a | For each group, the numbers of participants who were approached and/or assessed for eligibility, randomly assigned, received intended treatment, and were assessed for each objective | 9-10; Fig 1 |
|  | 13b | For each group, losses and exclusions after randomisation, together with reasons | 10 |
| Recruitment | 14a | Dates defining the periods of recruitment and follow-up | 6, 10 |
|  | 14b | Why the pilot trial ended or was stopped | Planned completion; 10 |
| Baseline data | 15 | A table showing baseline demographic and clinical characteristics for each group | 10-11 (Table 1) |
| Numbers analysed | 16 | For each objective, number of participants (denominator) included in each analysis. If relevant, these numbers  should be by randomised group | 12-13 |
| Outcomes and estimation | 17 | For each objective, results including expressions of uncertainty (such as 95% confidence interval) for any  estimates. If relevant, these results should be by randomised group | 12-13; S7-S9 |
| Ancillary analyses | 18 | Results of any other analyses performed that could be used to inform the future definitive trial | S5, S8-S9 |
| Harms | 19 | All important harms or unintended effects in each group (for specific guidance see CONSORT for harms) | 6, 11 |
|  | 19a | If relevant, other important unintended consequences | 15-16 |
| Discussion | | | |
| Limitations | 20 | Pilot trial limitations, addressing sources of potential bias and remaining uncertainty about feasibility | 17-18 |
| Generalisability | 21 | Generalisability (applicability) of pilot trial methods and findings to future definitive trial and other studies | 18-19 |
| Interpretation | 22 | Interpretation consistent with pilot trial objectives and findings, balancing potential benefits and harms, and  considering other relevant evidence | 16-19 |
|  | 22a | Implications for progression from pilot to future definitive trial, including any proposed amendments | 19 |
| Other information | | |  |
| Registration | 23 | Registration number for pilot trial and name of trial registry | 1, 4 |
| Protocol | 24 | Where the pilot trial protocol can be accessed, if available | 4 |
| Funding | 25 | Sources of funding and other support (such as supply of drugs), role of funders | Title Page |
|  | 26 | Ethical approval or approval by research review committee, confirmed with reference number | Title Page |

Citation: Eldridge SM, Chan CL, Campbell MJ, Bond CM, Hopewell S, Thabane L, et al. CONSORT 2010 statement: extension to randomised pilot and feasibility trials. BMJ. 2016;355. This is an Open Access article distributed in accordance with the terms of the Creative Commons Attribution (CC BY 3.0) license (<http://creativecommons.org/licenses/by/3.0/>), which permits others to distribute, remix, adapt and build upon this work, for commercial use, provided the original work is properly cited.

*We strongly recommend reading this statement in conjunction with the CONSORT 2010, extension to randomised pilot and feasibility trials, Explanation and Elaboration for important clarifications on all the items. If relevant, we also recommend reading CONSORT extensions for cluster randomised trials, non-inferiority and equivalence trials, non-pharmacological treatments, herbal interventions, and pragmatic trials. Additional extensions are forthcoming: for those and for up-to-date references relevant to this checklist, see [www.consort-statement.org](http://www.consort-statement.org)

**S1: Other Measures used in INACT**

**Primary Outcome Measures**

| **Outcome Measure** | **Measure Description** | **Time Frame** |
| --- | --- | --- |
| Intervention Feasibility (School Staff and Link Workers) | Feasibility of Intervention Measure (FIM) is assessed using 4 questions each on a five-point Likert scale (scoring between 4-20). Higher scores indicate higher intervention feasibility | 6 months |
| Intervention Acceptability (School Staff and Link Workers) | Acceptability of Intervention Measure (AIM) is assessed using 4 questions each on a five-point Likert scale (scoring between 4-20). Higher scores indicate higher intervention acceptability | 6 months |
| Intervention Appropriateness (School Staff and Link Workers) | Intervention Appropriateness Measure (IAM) is assessed using 4 questions each on a five-point Likert scale (scoring between 4-20). Higher scores indicate higher intervention appropriateness | 6 months |

**Secondary Outcome Measures**

| **Outcome Measure** | **Measure Description** | **Time Frame** |
| --- | --- | --- |
| Peer loneliness | LACA - Peer Subscale is assessed using 12 questions each on a four-point Likert scale (scoring between 12-48). Higher scores indicate higher peer loneliness | 3 and 6 months |
| Wellbeing | Kidscreen-52 is assessed using 6 questions each on a five-point Likert scale (scoring between 6-30). Higher scores indicate greater well-being. | 3 and 6 months |
| Mental health (emotional difficulties) | Me and My feelings is assessed using 10 questions on a 3-point Likert scale (scoring between 0-20). Higher scores indicate higher emotional difficulties | 3 and 6 months |
| Service Use | Client Service Receipt of Inventory is assessed using 11 questions on a five-point Likert scale. Scoring can be looked at by individual items (i.e. score between 1-5) or by scoring all items (i.e. scores between 11-55). Higher scores indicate more contact with a service/services. | 3 and 6 months |
| Stress | Perceived Stress Scale 4 is assessed using 4 questions on a five-point Likert scale (scoring between 0-16). Higher scores indicate higher levels of perceived stress | 3 and 6 months |
| Loneliness | Good Childhood Index is assessed using 3 questions on a three-point Likert scale (scoring between 3-9). Higher scores indicate higher reported loneliness. | 6 Months |

**Other Measures**

| **Outcome Measure** | **Measure Description** | **Time Frame** |
| --- | --- | --- |
| Family Support | Moderator using questions from the Student Resilience Survey which is assessed using 4 questions each on a five-point likert scale (scoring between 4-20). Higher scores indicate higher family support. | Baseline only |
| School Support | Moderator using questions from the Student Resilience Survey which is assessed using 4 questions each on a five-point Likert scale (scoring between 4-20). Higher scores indicate higher school support. | Baseline only |
| Social support | Moderator using CYRM-R which is assessed using 4 questions each on a five-point Likert scale (scoring between 4-20). Higher scores indicate higher social support | 3 and 6 months |
| Local Environment | Moderator using questions from the HBSC 2022 assessed using 4 questions each on a five-point Likert scale (scoring between 4-20). Higher scores indicate a more positive view of ones local environment | 3 and 6 months |
| Activity Engagement | Moderator using questions from the BeeWell survey which is assessed using 11 questions each on a six-point Likert scale (scoring between 11-66). Higher scores indicate higher daily engagement with more activities. Each item can also be scored individually and indicates more frequent engagement with that activity. | 3 and 6 months |
| Bullying | Moderator using kidscreen-52 (primary school pupils) which is assessed using 3 questions each on a five-point Likert scale (scoring between 3-15). Higher scores indicate higher levels of bullying | 3 and 6 months |
| Bullying | Moderator using the BeeWell survey (secondary school pupils) which is assessed using 3 questions each on a four-point Likert scale (scoring between 0-9). Higher scores indicate higher levels of bullying. | 3 and 6 months |
| Social structure and quality | Moderator using questions adapted from PISA 2022 and MCS4 which is assessed using five questions. One of which requires an open-ended numerical value and four questions which are rated on a five-point Likert scale (scoring between 0-16). Higher scores indicate a greater social structure and friendship quality. | 3 and 6 months |
| Problem Solving | Mechanism using the Student Resilience Survey which is assessed using 3 questions each on a five-point Likert scale (scoring between 3-15). Higher scores indicate higher problem solving with others | 3 and 6 months |
| Flow | Mechanism using the General Flow Proneness Scale (secondary school pupils only) which is assessed using 13 questions each on a five-point Likert scale (scoring between 13-65). Higher scores indicate higher flow experience | 3 and 6 months |
| Therapeutic Alliance | Mechanism using the Session Feedback Questionnaire which is assessed using 4 questions each on a five-point Likert scale (scoring between 4-20). Higher scores indicate higher therapeutic alliance. This is for individuals in the social prescribing intervention only | 3 and 6 months |

**S2: Interview Guides**

***S2a: Link Worker Topic Guide***

A. Professional background

1. Can you tell me a bit about your background and experience working as a social prescriber and with young people?

B. Delivery

1. Could you tell me about your experiences of delivering the social prescribing sessions to young people as part of this study?
2. Can you talk about a session that you felt went well?
3. Can you talk about a session that was more challenging for you?
4. Do you think any training or guidelines you had prior to the sessions were adequate to deal with such challenges? Why or why not?
5. Did you find that some young people engaged more with the sessions than others? Can you think of any reasons that may have contributed to differences in engagement?
6. Do you think that social prescribing worked better for some young people than others? If so, why?
7. Is there anything you might want to change about the delivery of the sessions to make them work better?
8. Could you tell me about your experiences of working with the school to deliver social prescribing for young people?
9. Could you tell me about your experiences of working with parents or guardians to deliver social prescribing for young people?

C. Perceived impact and unintended consequences

1. Could you describe the difficulties with loneliness or low community connection that the young people you worked with in this study were experiencing?
2. What are your views about using social prescribing to help young people who report low community connections or feel lonely?
3. What impact, if any, do you think your sessions had on the young people who took part?
4. Did your sessions have any other impacts that you were not expecting to see?

D. Intention to adopt, sustained use and research participation

1. How willing would you be to continue or adopt social prescribing for young people who report low community connections or feel lonely in the future?
2. Going forward, what might be the potential barriers or facilitators, if any, to the continued use or availability of social prescribing for these young people?
3. How did you find taking part in this study overall as a link worker?
4. How did you find the research procedures, such as interviews or data collection, as part of the project?

***S2b: YP Topic Guide***

A. Social and community support before intervention

1. Before you started meeting with your social prescriber, how were you feeling?
2. Thinking about before you met your social prescriber, what types of social and community activities were you involved in?
3. Could you tell me about why you decided to meet with your social prescriber?

B. Acceptability, feasibility and suitability of intervention

Meetings with the social prescriber

1. Could you tell me about your experience of meeting with the social prescriber?
2. Did you find it easy or difficult to attend the meeting(s) with the social prescriber?
3. Did meeting with the social prescriber help how you have been feeling at all?
4. Did you talk to your friends or family about meeting with the social prescriber?

Social and community support after intervention
5. Could you tell me about any activities or groups you tried out after meeting the social prescriber?

If they took part in activities:
6. Did you enjoy the activity?
7. Did you find it easy or difficult to attend the activity?
8. Did you find the activity helped how you have been feeling at all?

If they did not take part in any activities:
9. Could you tell me why you did not try out any community activities after meeting with the social prescriber?

C. Intention to adopt, sustained engagement and research participation

1. Since meeting your social prescriber, what types of social and community activities are you involved in?
2. Is there anything you have learnt from meeting with your social prescriber that you use, or plan to use in your everyday life?
3. [If they took part in an activity] Is there anything you have learnt from the activity that you use, or plan to use in your everyday life?
4. How did you find taking part in this research project overall?

**S3: Characteristics LWs participating in interviews**

| LW | Gender | Ethnicity | Site |
| --- | --- | --- | --- |
| 1 | Male | White/White British | Manchester |
| 2 | Non-binary | White/White British | London |
| 3 | Female | Did not disclose | Leeds |

**S4: Characteristics of YP participating in interviews**

| YP | Age | Gender | Ethnicity | Site | GCI score | SP engagement |
| --- | --- | --- | --- | --- | --- | --- |
| 1 | 9-13 | Girl | Asian / Asian British | London | Highest observed | High engagement (4–7 sessions) |
| 2 | 9-13 | Boy | White/White British | Leeds | Very elevated | Lower engagement (1–3 sessions) |
| 3 | 9-13 | Girl | Asian / Asian British | London | Elevated | High engagement (4–7 sessions) |
| 4 | 9-13 | Girl | Black / Black British | London | Elevated | High engagement (4–7 sessions) |
| 5 | 9-13 | Boy | Black / Black British | London | Elevated | High engagement (4–7 sessions) |

**S5: Preliminary power calculation**

The following calculation was used for the INACT pilot

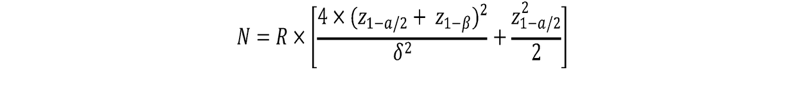

Where:
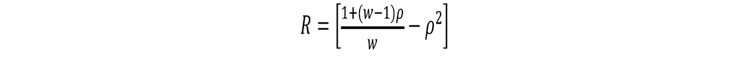

W is the number of post-intervention measures, which equals 3. We made the following assumptions: (1) the within-person correlation ($\rho$) was 0.6. (2) the standardised effect size ($\delta$) was 0.3^[1]^. We aimed to have 80% power ($\beta$) at the 5% significance level ($a$) for a two-sided test. This gives us a total sample size of 131. We estimated that each LW will see 25 participants and assumed a LW effect of 0.02. After adjusting for the intraclass correlation, we need a sample size of 194. With anticipated loss to follow up of 15% in both groups, we need a total sample size of 316 at baseline. Under realistic assumptions (ONS YP loneliness prevalence = 14%, participation rate=71%), we need to survey a minimum number of 3179 YPs to reach the targeted sample size (N=316). This calculation will be revised following pilot data. Using national data^43^, we have assumed that each primary school class has 25 participants, and each secondary school class has 20 participants. With 2 classes per each year group in years 4 and 5 (100 pupils per primary school) and 3 classes in each of Years 7 and 8 (120 pupils per secondary school). Five primary and secondary schools per city equals 3300 pupils

**S6: Stop-Go Criteria**

| Stop-go criteria |  |  |  |  |
| --- | --- | --- | --- | --- |
| Method | **Indicator** | **Fully met (proceed)** | **Partially met (proceed with amendments agreed by TSC)** | **Not met (stop)** |
| Acceptability of intervention | Qualitative reports of acceptability from young people, school staff and link workers | The majority of stakeholders report the social prescribing intervention is acceptable | Some stakeholders report the social prescribing intervention is not acceptable but identify alterations that need to occur | The majority of stakeholders report the social prescribing intervention is not acceptable |
|  | Proportion of young people attending an appointment with a link worker | 55-100% | 40-54% | 0-39% |
| Acceptability of evaluation framework | Qualitative reports of acceptability from young people, school staff and link workers | The majority of stakeholders report the social prescribing intervention is acceptable | Some stakeholders report the social prescribing intervention is not acceptable but identify alterations that need to occur | The majority of stakeholders report the social prescribing intervention is not acceptable |
|  | Proportion of young people completing baseline measures | 55-100% | 40-54% | 0-39% |
| Ability to collect 3-month data from pupils who completed baseline measures | Proportion of young people completing 3 month follow up | 55-100% | 40-54% | 0-39% |
| Negative consequences of the intervention | Reported negative/adverse effects of social prescribing from young people, school staff or link workers | No evidence of substantially negative effects on young people | - | Evidence of substantially negative effects on young people |

**S7: Main loneliness model**

***Table S7. Results from Bayesian growth curve models on loneliness***

|  | Response scale | | | Standardised scale | | |
| --- | --- | --- | --- | --- | --- | --- |
|  | Estimate | 95% CI | Rhat | Estimate | 95% CI | Rhat |
| Intercept | 7.60 | [7.36, 7.83] | 1.00 | 0.68 | [0.54, 0.81] | 1.00 |
| 3-month | -1.83 | [-2.45, -1.22] | 1.00 | -1.04 | [-1.39, -0.68] | 1.00 |
| 6-month | -2.12 | [-2.79, -1.46] | 1.00 | -1.20 | [-1.57, -0.82] | 1.00 |
| sp | 0.11 | [-0.23, 0.46] | 1.00 | 0.06 | [-0.14, 0.26] | 1.00 |
| 3-month:sp | -0.57 | [-1.46, 0.33] | 1.00 | -0.32 | [-0.81, 0.17] | 1.00 |
| 6-month:sp | 0.20 | [-0.75, 1.11] | 1.00 | 0.11 | [-0.42, 0.65] | 1.00 |
| SD (Intercept) | 0.30 | [0.02, 0.70] | 1.00 | 0.18 | [0.01, 0.42] | 1.01 |
| SD (3-month) | 1.53 | [1.15, 1.92] | 1.00 | 0.87 | [0.66, 1.11] | 1.00 |
| SD (6-month) | 1.55 | [1.15, 2.00] | 1.00 | 0.89 | [0.66, 1.15] | 1.00 |
| Cor (Intercept, 3-month) | 0.18 | [-0.54, 0.88] | 1.01 | 0.16 | [-0.53, 0.88] | 1.00 |
| Cor (Intercept, 6-month) | 0.33 | [-0.41, 0.93] | 1.01 | 0.28 | [-0.42, 0.90] | 1.01 |
| Cor (3-month, 6-month) | 0.48 | [0.16, 0.74] | 1.00 | 0.48 | [0.17, 0.74] | 1.00 |
| Sigma | 0.72 | [0.41, 0.90] | 1.00 | 0.40 | [0.18, 0.51] | 1.01 |

**S8:** **Peer loneliness model**

***Table S8. Results from Bayesian growth curve models on peer loneliness***

|  | Response scale | | | Standardised scale | | |
| --- | --- | --- | --- | --- | --- | --- |
|  | Estimate | 95% CI | Rhat | Estimate | 95% CI | Rhat |
| Intercept | 32.43 | [30.4, 34.47] | 1.00 | 0.45 | [0.21, 0.71] | 1.00 |
| 3-month | -5.43 | [-8.23, -2.7] | 1.00 | -0.67 | [-1.01, -0.33] | 1.00 |
| 6-month | -5.73 | [-8.73, -2.87] | 1.00 | -0.71 | [-1.06, -0.35] | 1.00 |
| sp | 0.58 | [-2.39, 3.52] | 1.00 | 0.07 | [-0.28, 0.44] | 1.00 |
| 3-month:sp | -2.05 | [-6.07, 1.89] | 1.00 | -0.26 | [-0.75, 0.22] | 1.00 |
| 6-month:sp | -0.98 | [-5.00, 3.40] | 1.00 | -0.12 | [-0.63, 0.38] | 1.00 |
| SD (Intercept) | 4.46 | [2.67, 6.52] | 1.01 | 0.55 | [0.34, 0.79] | 1.00 |
| SD (3-month) | 3.84 | [0.30, 8.12] | 1.02 | 0.50 | [0.06, 0.96] | 1.01 |
| SD (6-month) | 3.72 | [0.33, 8.28] | 1.02 | 0.48 | [0.04, 0.99] | 1.01 |
| Cor (Intercept, 3-month) | 0.16 | [-0.52, 0.89] | 1.01 | 0.12 | [-0.52, 0.88] | 1.01 |
| Cor (Intercept, 6-month) | 0.10 | [-0.58, 0.88] | 1.01 | 0.06 | [-0.58, 0.84] | 1.01 |
| Cor (3-month, 6-month) | 0.60 | [-0.45, 0.98] | 1.00 | 0.62 | [-0.48, 0.98] | 1.00 |
| Sigma | 5.11 | [2.38, 6.27] | 1.02 | 0.63 | [0.40, 0.77] | 1.01 |

**S9: Sensitivity analyses**

***Table S9a. Results from Bayesian growth curve models on loneliness excluding participants in the SP group who did not receive SP***

|  | Response scale | | | Standardised scale | | |
| --- | --- | --- | --- | --- | --- | --- |
|  | Estimate | 95% CI | Rhat | Estimate | 95% CI | Rhat |
| Intercept | 7.60 | [7.37, 7.83] | 1.00 | 0.68 | [0.54, 0.81] | 1.00 |
| 3-month | -1.84 | [-2.44, -1.24] | 1.00 | -1.03 | [-1.38, -0.68] | 1.00 |
| 6-month | -2.12 | [-2.74, -1.47] | 1.00 | -1.19 | [-1.56, -0.81] | 1.00 |
| sp | 0.05 | [-0.30, 0.40] | 1.00 | 0.03 | [-0.17, 0.23] | 1.00 |
| 3-month:sp | -0.44 | [-1.34, 0.39] | 1.00 | -0.26 | [-0.77, 0.26] | 1.00 |
| 6-month:sp | 0.13 | [-0.85, 1.10] | 1.00 | 0.07 | [-0.51, 0.61] | 1.00 |
| SD (Intercept) | 0.26 | [0.02, 0.66] | 1.01 | 0.16 | [0.01, 0.39] | 1.01 |
| SD (3-month) | 1.51 | [1.14, 1.92] | 1.00 | 0.86 | [0.65, 1.10] | 1.00 |
| SD (6-month) | 1.57 | [1.17, 2.03] | 1.00 | 0.89 | [0.66, 1.15] | 1.00 |
| Cor (Intercept, 3-month) | 0.22 | [-0.63, 0.90] | 1.02 | 0.24 | [-0.48, 0.89] | 1.00 |
| Cor (Intercept, 6-month) | 0.16 | [-0.63, 0.88] | 1.02 | 0.18 | [-0.52, 0.87] | 1.00 |
| Cor (3-month, 6-month) | 0.56 | [0.26, 0.80] | 1.00 | 0.56 | [0.26, 0.80] | 1.00 |
| Sigma | 0.72 | [0.43, 0.89] | 1.01 | 0.40 | [0.22, 0.51] | 1.01 |

***Table S9b. Results from Bayesian growth curve models on loneliness excluding participants in the SP group who did not receive SP and participants who started intervention later***

|  | Response scale | | | Standardised scale | | |
| --- | --- | --- | --- | --- | --- | --- |
|  | Estimate | 95% CI | Rhat | Estimate | 95% CI | Rhat |
| Intercept | 7.60 | [7.37, 7.83] | 1.00 | 0.68 | [0.55, 0.81] | 1.00 |
| 3-month | -1.83 | [-2.42, -1.21] | 1.00 | -1.04 | [-1.40, -0.69] | 1.00 |
| 6-month | -2.10 | [-2.78, -1.42] | 1.00 | -1.20 | [-1.57, -0.84] | 1.00 |
| sp | 0.07 | [-0.32, 0.45] | 1.00 | 0.04 | [-0.19, 0.25] | 1.00 |
| 3-month:sp | -0.45 | [-1.40, 0.48] | 1.00 | -0.24 | [-0.78, 0.31] | 1.00 |
| 6-month:sp | -0.10 | [-1.15, 0.91] | 1.00 | -0.05 | [-0.65, 0.54] | 1.00 |
| SD (Intercept) | 0.33 | [0.03, 0.74] | 1.01 | 0.18 | [0.01, 0.41] | 1.01 |
| SD (3-month) | 1.51 | [1.12, 1.92] | 1.00 | 0.86 | [0.64, 1.10] | 1.00 |
| SD (6-month) | 1.60 | [1.18, 2.09] | 1.00 | 0.91 | [0.66, 1.21] | 1.00 |
| Cor (Intercept, 3-month) | 0.30 | [-0.43, 0.92] | 1.01 | 0.32 | [-0.39, 0.92] | 1.01 |
| Cor (Intercept, 6-month) | 0.23 | [-0.50, 0.89] | 1.00 | 0.26 | [-0.47, 0.91] | 1.00 |
| Cor (3-month, 6-month) | 0.62 | [0.32, 0.86] | 1.00 | 0.63 | [0.31, 0.85] | 1.00 |
| Sigma | 0.69 | [0.31, 0.88] | 1.02 | 0.40 | [0.19, 0.54] | 1.01 |

1. Overall effect size identified in Eccles and Qualter 2021 loneliness review
